# A Two-Stage Plasma Biomarker Algorithm Reduces Amyloid PET Use and the p-Tau217 Gray Zone

**DOI:** 10.64898/2026.02.03.26345302

**Authors:** Harthik Parankusham, Casey Vanderlip, Colin Birkenbihl, Eashwar Krishna, Chizobam Ugboaja, Andrew Budson, Brandon Frank, Alzheimer’s Disease Neuroimaging Initiative

**Affiliations:** Department of Physiology & Neurobiology, UConn Computational Biology Core, University of Connecticut, Storrs, CT 06269-3125, USA; Department of Neurobiology and Behavior, 1424 Biological Sciences III Irvine, University of California Irvine, Irvine, CA, 92697 USA; Department of Neurology, Massachusetts General Hospital, Harvard Medical School, Boston, MA 02115, USA; Department of Molecular & Cell Biology, University of Connecticut, 91 North Eagleville Road, Unit 3125, Storrs, CT 06269-3125, USA; Yale School of Medicine, Yale New Haven Health, 333 Cedar Street, New Haven, CT 06510, USA; VA Boston Healthcare System, Boston, MA, USA; Alzheimer’s Disease Research Center, Boston University Chobanian & Avedisian School of Medicine, Boston, MA, USA

**Author notes:** Corresponding author. (H.P). **Alzheimer’s Disease Neuroimaging Initiative:** Data used in the preparation of this article were obtained from the Alzheimer’s Disease Neuroimaging Initiative (ADNI) database (adni.loni.usc.edu). As such, the investigators within the ADNI contributed to the design and implementation of ADNI and/or provided data but did not participate in analysis or writing of this report. A complete listing of ADNI investigators can be found at: http://adni.loni.usc.edu/wp-content/uploads/how_to_apply/ADNI_Acknowledgement_List.pdf.

**Keywords:** p-Tau217, Alzheimer’s disease, cognitively unimpaired, amyloid PET, gray zone

## Abstract

**Objective:** Plasma p-Tau217 leaves 30-50% of patients in an indeterminate “gray zone” often requiring Aβ-PET. We developed GRAD (Gatekeeper-Reflex for Alzheimer’s Disease), a two-stage Aβ+ prediction model to improve classification while reserving Aβ-PET for residual uncertainty.

**Methods:** GRAD was developed in 145 cognitively impaired ADNI participants and externally validated in 1,644 cognitively unimpaired A4/LEARN participants. Stage 1 used p-Tau217 alone; probabilities between 0.25-0.75 were considered indeterminate. Stage 2 used L2-regularized logistic regression combining plasma-biomarkers, genetic and demographic features. Aβ-PET served as the reference standard (ADNI florbetapir SUVR>1.11; A4 Centiloid≥20).

**Results:** Full model AUC was 0.915 (95% CI, 0.859-0.961) internally and 0.879 (95% CI, 0.861-0.895) externally. In the external gray zone (*n*=630), Stage 2 improved discrimination over p-Tau217 alone (AUC 0.793 vs 0.719; P<.001), increased correct classification without PET from 41.4% to 56.2% and reduced PET referral from 35.9% to 24.3%. In the p-Tau181 subset (*n*=256), Stage 2 outperformed p-Tau181 (AUC 0.799 vs 0.597; P<.001). Overall, 89.0% of ADNI and 90.7% of A4/LEARN participants were classified without PET; modeled testing costs were 44-52% lower than p-Tau217 screening with PET confirmation of the gray zone.

**Interpretation:** Reflex multimarker testing improves indeterminate p-Tau217 classification while retaining Aβ-PET for residual uncertainty. GRAD may reduce PET use and testing costs in memory-clinic triage, prevention-trial enrichment, and disease-modifying therapy workup.

## 1. Introduction

Blood-based biomarkers (BBMs) for Alzheimer’s disease (AD) enable detection of cerebral amyloid-β (Aβ) pathology without the cost and limited accessibility of Aβ-positron emission tomography (Aβ-PET) or the invasiveness of cerebrospinal fluid (CSF) analysis^1–3^. Recently, phosphorylated tau217 (p-Tau217) has exhibited the highest accuracy in plasma for identifying Aβ positive (Aβ+) individuals^4–10^, even in the earliest stages of disease^11, 12^. In clinical practice, p-Tau217 plays the crucial role of screening individual eligibility for prevention studies and recent FDA-approved disease-modifying therapies (DMTs)^3, 13^.

However, for preclinical-AD and DMT-eligible individuals where treatment decisions are critical, there exists a large overlap of p-Tau217 values in classifying Aβ+ versus Aβ-^13, 14^. As a result ∼30-50% of individuals fall within a “gray zone” where p-Tau217 alone cannot confidently classify Aβ-status^5, 13–15^. Several studies have attempted to resolve this uncertainty, including staged p-Tau217/Aβ-PET workflows and complementary BBMs such as p-Tau181^15–20^. However, these studies had modest sample sizes, lacked external validation, and did not specifically target MCI and preclinical AD - the populations most likely to benefit from currently available DMTs. Although multimarker approaches incorporating p-Tau217, Aβ42/40^21–24^, and glial fibrillary acidic protein (GFAP) show promise^25, 26^, they have not yet been adequately tested within the gray zone population.

We hypothesized that a plasma-based staged model - similar to reflex testing in laboratory medicine (*53,54*) - could resolve gray zone uncertainty in DMT-eligible and preclinical-AD individuals. First, a p-Tau217 screen would identify clear-cut cases, while a machine-learning (ML) classification model integrating several plasma and demographic features would address uncertain cases. Only truly uncertain cases would require expensive, but confirmatory Aβ-PET scans.

In this study, we present the Gatekeeper-Reflex model for AD (GRAD), a two-stage plasma-based algorithm to classify individuals in the gray zone for Aβ-positivity, with the goal of reducing residual confirmatory Aβ-PET scans. We aimed to: (i) assess GRAD internal and external performance in DMT-eligible and pre-clinical populations (ii) benchmark GRAD against existing methods to ameliorate the gray zone and (iii) simulate our model’s health-economic impact.

## 2. Methods

### 2.1 Study Populations

We drew participants from two independent cohorts for model development and external validation. GRAD was developed in impaired participants and externally validated in preclinical-AD individuals, thus ensuring model generalizability across clinically distinct populations.

#### 2.1.1 Development and Training Cohort

We selected cognitively impaired participants from the Alzheimer’s Disease Neuroimaging Initiative (ADNI) (*31*) whose plasma p-Tau217 and Aβ-PET were collected at the same time. The 145 individuals who met these criteria, formed the development cohort: MCI (n=117, 80.7%) and AD dementia (n=28, 19.3%). We considered ADNI florbetapir SUVR>1.11^36–38^ as Aβ+.

#### 2.1.2 Independent Validation Cohort

For the external validation cohort, we selected 1,644 participants with baseline p-Tau217 and Aβ status available from the Anti-Amyloid Treatment in Asymptomatic Alzheimer’s disease (A4) Study (1,119 in A4, 525 in LEARN) ^32–35^. This included 1,145 Aβ-positive and 499 Aβ-negative participants. We considered Centiloid≥20 as Aβ+ per the Centiloid standardization framework^33, 39^.

### 2.2 Plasma Assays and Biomarker Acquisition

ADNI plasma p-Tau217 was measured on two platforms: the Lumipulse G p-Tau217 assay (Fujirebio) at the University of Pennsylvania (UPENN) in 81 of the 145 ADNI participants, and the Janssen p-Tau217 assay in the remaining 64^9^. Plasma samples were collected and stored as per ADNI biofluid protocols^31^. The UPENN panel provided GFAP in 81 participants, and Aβ42/40 in 80. The Janssen subset contributed p-Tau217 only. The APOE genotype was available in 132 ADNI participants. APOE genotype was obtained from the ADNI APOE results file, and demographic data, diagnostic classification and florbetapir amyloid PET (AV45 SUVR) from ADNIMERGE. We also report analyte availability by cohort and platform (Table S1). All files were downloaded from the ADNI database (http://adni.loni.usc.edu/) under an approved data use agreement.

In A4, all 1,644 participants had p-Tau217 measured via the Lilly Research Laboratories MSD immunoassay^40^. GFAP and Aβ42/40 were measured on the Roche Elecsys platform in a subset of the cohort: GFAP in 931 participants, and Aβ42/40 in 919, with APOE genotype available in 1,642. Aβ42/40 was available to 474 A4-arm participants and 445 LEARN participants.

Participants missing an analyte were retained with the missing value imputed from the training-set median (Section 2.3), so the denominators reported throughout include them. We ensured test-set values were never imputed from the test cohort’s own distribution. We include a complete-panel cohort sensitivity analysis with no imputation performed (Table S2).

### 2.3 Data Pre-Processing and Harmonization

For pre-processing and harmonization, we Z-normalized all biomarkers measured from blood plasma to a pre-specified reference population of CU, Aβ-negative individuals within each cohort and assay platform^8, 9^ (Table S3). The raw biomarker values were log-transformed and Z-scores were computed as: Z = (log(1+x) - μ_ref) / σ_ref. This preprocessing step was crucial since both cohorts utilized different assays for p-Tau217 (Table S1).

The Aβ42/40 ratio was the only exception and entered our model as a raw log ratio. Since it is a ratio of two analytes measured on the same platform from the same sample, the platform scale factors cancel within it. Anchoring it to the reference subset therefore adds noise without removing bias.

### 2.4 Design of the Two-Stage Algorithm

The model consisted of two stages. Stage 1 (Gatekeeper) was a univariate logistic regression model using only p-Tau217 to predict Aβ+ individuals. We set classification thresholds based on p-Tau217 predictive confidence: Aβ-negative where P<0.25 and Aβ-positive where P>0.75. Cases falling within the intermediate gray zone (0.25 ≤P≤0.75)^15^ were triaged to the next stage (Figure 2A). In Stage 2 (Reflex), for gray zone cases, we implemented an L2-regularized logistic regression (C = 0.1 set apriori to promote strong shrinkage, balanced class weights) on features standardized within each training fold (StandardScaler, scikit-learn). The feature set comprised p-Tau217, GFAP, a tau-Aβ divergence ratio (log[p-Tau217] - log[Aβ42/40]), log Aβ42/40, a GFAP by p-Tau217 interaction term, age, and APOE ε4 carrier status (Figure 1).We manually selected BBM interaction terms based on biological priors, instead of utilizing ML model-based discovery which could create cohort dependence.

**Figure 1.**
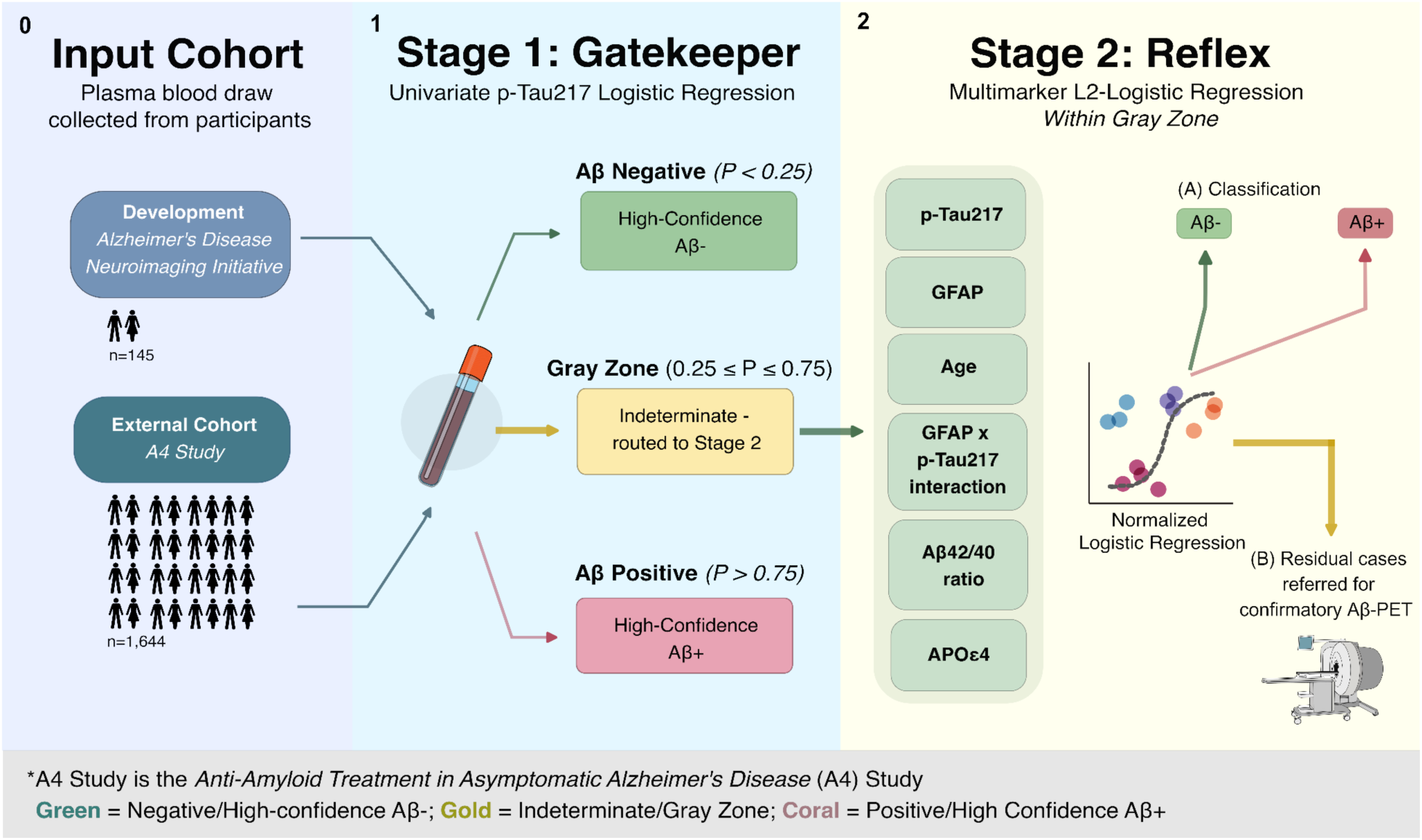
Study Design and Workflow. Participants from the development cohort (ADNI, n=145 cognitively impaired) and the external validation cohort (A4 and LEARN, n=1,644 cognitively unimpaired) contributed a single plasma draw. In Stage 1 (Gatekeeper), a univariate logistic regression on harmonized p-Tau217 returns a predicted probability of Aβ positivity: participants below 0.25 are classified high-confidence Aβ-negative (green), those above 0.75 high-confidence Aβ-positive (coral), and those between 0.25 and 0.75 fall in the indeterminate gray zone (gold) and are triaged forward.

**Figure 2.**
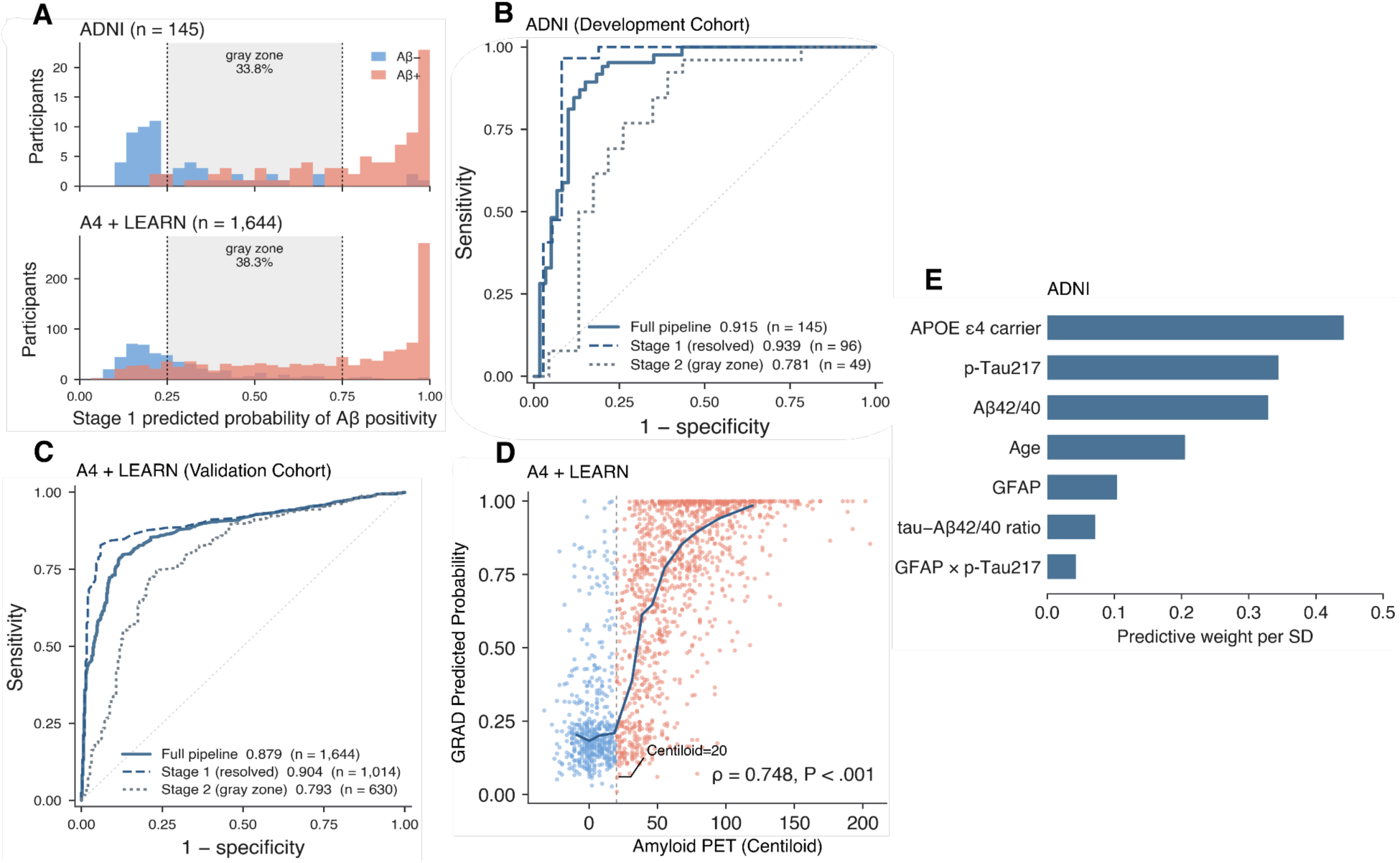
Model Cross-Cohort Performance. **(A)** Distribution of Stage 1 predicted probability of Aβ positivity in ADNI (top, n=145) and A4 + LEARN (bottom, n=1,644), stratified by Aβ-PET status. Shading marks the 0.25-0.75 gray zone, which contained 33.8% of ADNI and 38.3% of A4 + LEARN participants. **(B)** Receiver operating characteristic curves in the ADNI development cohort under leave-one-out cross-validation: full pipeline (AUC 0.915, n=145), Stage 1 among resolved cases (0.939, n=96), and Stage 2 within the gray zone (0.781, n=49). **(C)** The same curves in the A4 + LEARN validation cohort, scored once by the ADNI-fitted model without refitting: full pipeline (0.879, n=1,644), Stage 1 (0.904, n=1,014), Stage 2 (0.793, n=630). **(D)** GRAD predicted probability against Aβ-PET Centiloid value in A4 + LEARN, colored by Aβ-PET status; the dashed line marks the Centiloid 20 positivity threshold and the solid line is a running median. **(E)** Ranked predictive weights of each feature and interaction term in Reflex (Stage 2) of ADNI.

We considered, in both cohorts, reflex cases with predicted probability outside 0.40-0.60 resolved, while cases within 0.40-0.60 remained indeterminate and referred for confirmatory Aβ-PET.

### 2.5 Model Validation Strategy

We internally evaluated our model using leave-one-out cross-validation (LOOCV). Data pre-processing and harmonization parameters were recalculated within each fold, and Stage 1 and Stage 2 were refit within each fold. We calculated percentile bootstrap confidence intervals (CIs; 2,000 resamples of the LOOCV predictions) for AUC, sensitivity, and specificity. After harmonizing platforms, we externally validated the ADNI-trained model on the independent A4 trial data. Importantly, the A4 data was scored once by the fitted model, and was never used for training, feature selection, or any thresholding choices.

For A4 external validation (A4-arm and LEARN), we performed a standard binary classification. We additionally derived a Spearman correlation between individual-level predicted probabilities and Aβ-PET centiloid values for a continuous validity check. We assessed calibration by the slope and intercept of observed outcome on the predicted log-odds.

GRAD’s statistical analysis consisted of AUC with bootstrap confidence intervals (2,000 resamples), sensitivity, specificity, positive predictive value (PPV), negative predictive value (NPV), likelihood ratios (LR+, LR-) under STARD 2015 diagnostic guidelines^41^, and Brier scores for calibration of probabilistic predictions. We only used DeLong’s test^42^ for paired comparisons of two models scored on the same participants. Comparing classification accuracy between p-Tau217 alone and GRAD Stage 2, evaluated on the same participants under the same indeterminate band, used McNemar’s test.

### 2.6 Benchmarking Against p-Tau217 and p-Tau181

We benchmarked GRAD Stage 2, first against p-Tau217 alone in the gray zone, then against p-Tau181 integration - the single published BBM-only workflow designed to address this problem^16^. Against p-Tau217, in both cohorts, we compared gray zone AUC by DeLong’s test, and applied the 0.40-0.60 indeterminate band to both scores - Stage 2 versus p-Tau217 - to compare the proportions determined correctly or incorrectly, and referred for PET. The paired reclassifications were assessed using McNemar’s test. Against p-Tau181, we assessed the same quantities in the 256 A4 gray zone participants with a p-Tau181 measurement. Two considerations apply in our benchmarking analysis. First, benchmarking Stage 2 versus p-Tau181 was limited to the preclinical A4 cohort, since only 17 of the 145 ADNI participants contained visit-matched p-Tau181 metrics. Second, we applied the same validation rule as the original p-Tau181 integration (Giacomucci et al.) study: A dichotomous cutoff derived by Youden’s index in the full cohort of impaired individuals. This meant no cases were referred to Aβ-PET, while GRAD abstains from classifying significantly low-confidence cases. Thus, we provide decision-level metrics alongside AUC.

### 2.7 Cost Impact Simulation

To quantify GRAD’s health economic impact, we developed a clinical decision-based model comparing three diagnostic strategies for a simulated cohort of 10,000 patients: Universal Aβ-PET (estimated conservatively $3,000 per scan), p-Tau217 screening + PET for gray zone cases ($350 per single-analyte p-Tau217 test + PET for all indeterminate cases), and our GRAD staged algorithm. GRAD is billed as a reflex assay: every patient receives the $350 p-Tau217 test, the multi-analyte add-on panel ($250) is charged only to the fraction routed to Stage 2, and PET is reserved for the residual indeterminate cases. Resolution rates were derived from the observed LOOCV and A4 validation results (Sections 3.2, 3.3, and 3.5). We determined unit costs based on 2024 U.S. Medicare reimbursement schedules^43–45^ and, where necessary, we assumed higher costs to avoid potential overestimation of savings (Table S4). We report cost per correct diagnosis, counting cases referred for PET as correctly diagnosed.

## 3. Results

### 3.1 Baseline Demographic Characteristics

The 145 participants from the ADNI cohort used for model development had a mean age of 73.3 ± 7.1 years, were 37.2% female, and 89.0% White (Table 1). 46.2% of participants with an available genotype were APOE ε4 carriers and the median p-Tau217 level was 0.139 pg/mL, with 58.6% Aβ positive. A4/LEARN participants had a mean age 71.5 ± 4.7 years, was 60.2% female; 47.4% were APOE ε4 carriers, the median p-Tau217 level was 0.192 pg/mL, and 69.6% were Aβ positive (Table 1).

**Table 1.** Baseline cohort distribution and characteristics.

| Characteristic | ADNI development cohort<br>(n = 145) | A4 + LEARN validation cohort<br>(n = 1,644) |
| --- | --- | --- |
| Age, years, mean (SD) | 73.3 (7.1) | 71.5 (4.7) |
| Female, n (%) | 54 (37.2) | 990 (60.2) |
| Education, years, mean (SD) | 15.8 (2.7) | 16.6 (2.7) |
| White, n (%) | 129 (89.0) | 1,546 (94.0) |
| MMSE, mean (SD) | 26.5 (3.3) | 28.8 (1.2) |
| APOE ε4 carrier, n (%)<sup>a</sup> | 61 (46.2) | 779 (47.4) |
| **Clinical status, n (%)** |  |  |
| Cognitively unimpaired | 0 | 1,644 (100) |
| Mild cognitive impairment | 117 (80.7) | 0 |
| AD dementia | 28 (19.3) | 0 |
| **Aβ status** |  |  |
| Aβ-PET positive, n (%) | 85 (58.6) | 1,145 (69.6) |
| Positivity threshold | Florbetapir SUVR > 1.11 | Centiloid ≥ 20 |
| **Plasma p-Tau217** |  |  |
| **Median (IQR), pg/mL** | 0.139 (0.067-0.356) | 0.192 (0.142-0.276) |
| **Assay platform** | Lumipulse G, n = 81;<br>Janssen, n = 64 | Lilly Research Laboratories MSD |
| **A4 randomized arms** | - | 1,119 (68.1) |
| **LEARN** | - | 525 (31.9) |
| **Panel analyte availability, n (%)** |  |  |
| **GFAP** | 81 (55.9) | 931 (56.6) |
| **Aβ42/40** | 80 (55.2) | 919 (55.9) |

Data are mean (SD), median (IQR), or n (%) unless otherwise indicated.
<sup>a</sup> Percentages are calculated among participants with available APOE genotype.
**Abbreviations:** AD, Alzheimer's disease; Aβ , amyloid-β ; GFAP, glial fibrillary acidic protein; IQR, interquartile range; MMSE, Mini-Mental State Examination; MSD, Meso Scale Discovery; PET, positron emission tomography; SUVR, standardized uptake value ratio.

### 3.2 Full Model Performance

In ADNI, the complete GRAD model achieved an AUC of 0.915 (95% CI, 0.859-0.961), accuracy of 87.6% (95% CI, 82.1%-93.1%), 89.4% sensitivity (95% CI, 82.7%-95.6%), 85.0% specificity (95% CI, 75.0%-93.0%), PPV 89.4%, NPV 85.0%, LR+ 5.96, LR-0.12, and a Brier score of 0.108 (Table S5). The model correctly classified 127 of 145 participants, with 76 true positives and 51 true negatives. Performance was consistent across cognitive status (MCI 0.906, 95% CI 0.839-0.958; AD dementia 0.958, 0.861-1.000), APOE ε4 status (ε4-0.921, 0.851-0.979; ε4+ 0.817, 0.624-0.978), sex (male 0.940, 0.882-0.983; female 0.865, 0.726-0.974), and age tertiles (0.853-0.969) (Figure S1). At the 90% sensitivity operating threshold (P<0.451), the model achieved 91.8% sensitivity with 81.7% specificity (NPV 87.5%, LR-0.10); at the 90% specificity threshold (P>0.667), 90.0% specificity with 81.2% sensitivity (PPV 92.0%, LR+ 8.12) (Table S6).

In A4 and LEARN, the ADNI-trained model achieved AUC 0.879 (95% CI, 0.861-0.895), with 76.7% sensitivity, 88.6% specificity, PPV 93.9% and NPV 62.3%. Subgroup performance in the external cohort was similarly stable (Figure 3A). Individual-level predicted probabilities correlated with Aβ-PET centiloid values (Spearman ρ=0.748, P<.001), indicating that model outputs track continuous amyloid burden beyond binary classification. Across the full pipeline, 89.0% of ADNI and 90.7% of A4 participants received a definitive Aβ classification without imaging.

**Figure 3.**
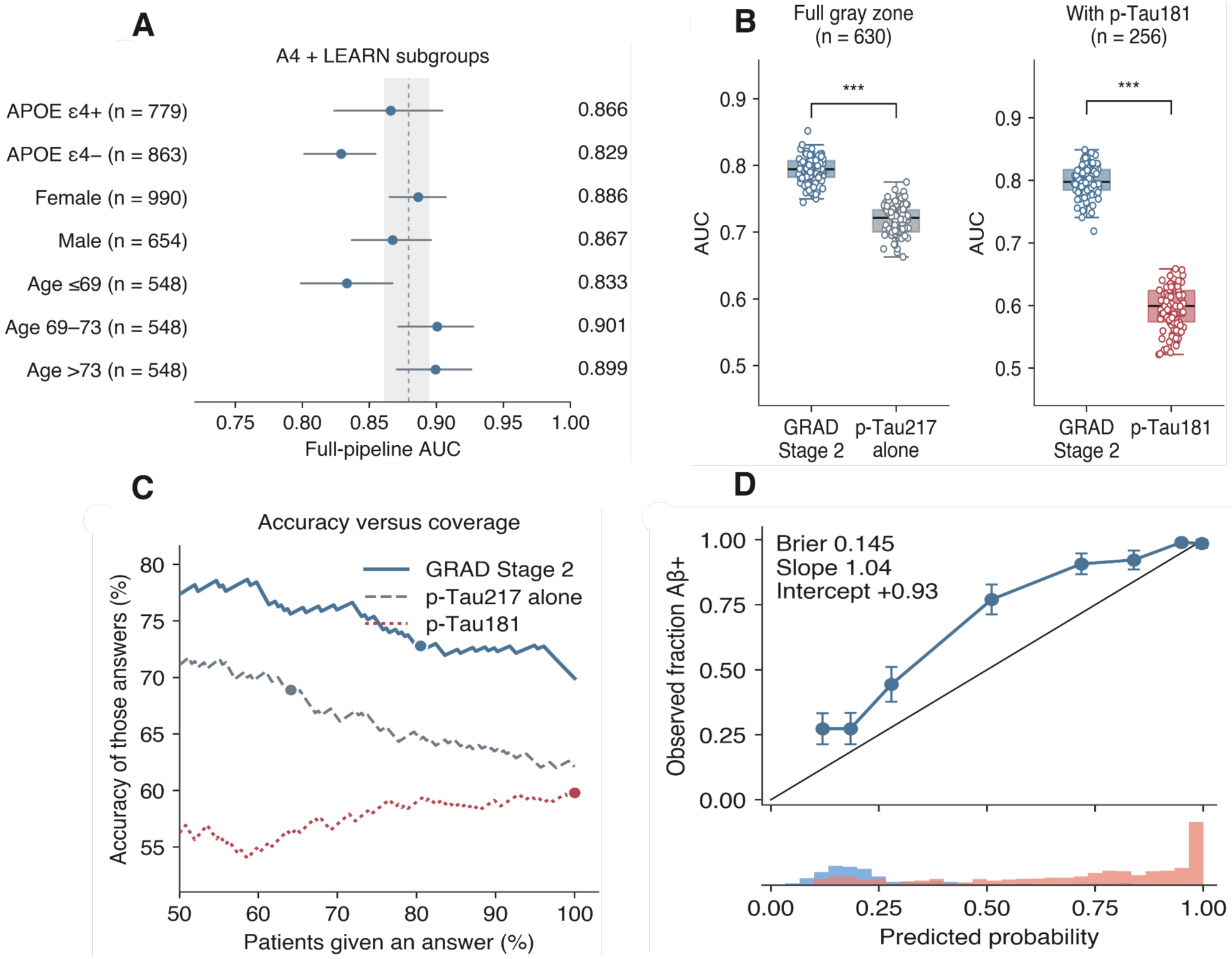
External Validation and Benchmarking Analysis. **(A)** Full-pipeline AUC across clinical subgroups of the A4 + LEARN validation cohort, with 95% bootstrap confidence intervals; the dashed line and shaded band show the whole-cohort estimate. **(B)** Bootstrap distributions of gray zone AUC. Left: GRAD Stage 2 versus p-Tau217 alone across the full external gray zone. Right: GRAD Stage 2 versus p-Tau181 in the subset with a visit-matched p-Tau181 measurement. Boxes show the interquartile range, the horizontal line the median. ***P < .001. **(C)** Accuracy against coverage in the external gray zone. Note as the low-confidence band is widened and fewer patients are given an answer, accuracy among those answered rises. GRAD Stage 2 and p-Tau217 alone under the 0.40-0.60 indeterminate band, and p-Tau181 at 100% coverage (Youden cutoff). **(D)** Calibration of the full pipeline in A4 + LEARN. Observed versus predicted Aβ positivity by decile of predicted probability with 95% confidence intervals; diagonal is perfect calibration. The lowest panel is the distribution of predicted probabilities by Aβ-PET status.

While discrimination and routing transferred, calibration, in large, did not. The calibration slope was 1.04 and the intercept +0.93 with mean predicted risk 57.5% against an observed prevalence of 69.6% (Brier 0.145) (Figure 3D).

### 3.3 Stage 1 Performance

In ADNI, using only the Gatekeeper model, 96 of the 145 participants (66.2%) placed outside the gray zone and were resolved with an AUC of 0.939 (95% CI, 0.866-0.995). 37 cases were classified as Aβ negative (NPV 91.9%), 59 cases were classified as Aβ positive (PPV 94.9%), for an accuracy of 93.8% among resolved cases, and 49 cases were in the gray zone (33.8%) which were routed to Stage 2.

In A4 and LEARN, the same model resolved 1,014 of 1,644 participants (61.7%) with an AUC of 0.904, classifying 408 as Aβ negative (NPV 70.1%) and 606 as Aβ positive (PPV 97.0%), for an accuracy of 86.2% among resolved cases and 630 (38.3%) entered the gray zone.

The fitted Stage 1 model was logit(P) = -1.023 + 1.124 × p-Tau217_Z, so the routing thresholds correspond to fixed harmonized p-Tau217 values and can be applied to a new patient by hand.

### 3.4 Stage 2 Performance

In ADNI, within the gray zone (n=49) (Figure 2A), Stage 2 achieved an AUC of 0.781 (95% CI, 0.630-0.913) with 75.5% accuracy, 76.9% sensitivity and 73.9% specificity, against 0.739 (0.592-0.867) for p-Tau217 alone on the same participants (+0.042; DeLong P = .54) (Figure 2B). In A4 and LEARN, within the gray zone (n=630) (Figure 2A), Stage 2 achieved an AUC of 0.793 (95% CI, 0.753-0.832) with 70.8% accuracy, 66.7% sensitivity and 80.0% specificity (Figure 2C), against 0.719 (0.675-0.762) for p-Tau217 alone (+0.073) -both comparisons are reported in full in Section 3.5.

We found that discrimination inside the band rests mainly on three predictors: APOE ε4 carrier status (odds ratio per SD 3.14; 95% CI, 1.53-6.44; P = .002), p-Tau217 (2.34; 1.20-4.56; P = .013) and Aβ42/40 (0.32; 0.13-0.77; P = .011). GFAP (0.80; P = .48) and the GFAP by p-Tau217 interaction (0.94; P = .83) contribute little (Figure 2E; Table S7). Drop-one ablation in the external cohort gives the same ordering: removing APOE ε4 costs 0.034 AUC and Aβ42/40 costs 0.030, while removing GFAP, age or the interaction did not significantly degrade performance (Table S8).

Stage 2 resolved 33 of the 49 ADNI gray zone cases (67.3%) and 477 of the 630 in A4 (75.7%), leaving 16 (11.0% of the development cohort) and 153 (9.3% of the validation cohort) indeterminate and referred for confirmatory Aβ-PET.

### 3.5 Benchmarking Against p-Tau217 and p-Tau181

Against p-Tau217 alone, Stage 2 improved gray zone discrimination by 0.042 in ADNI (0.739 to 0.781; DeLong P = .54, n=49) and by 0.073 in A4 (0.719 to 0.793; DeLong P < .001, n=630). Applying the identical 0.40-0.60 indeterminate band to both scores in the external cohort, p-Tau217 alone resolved 41.4% of gray zone patients correctly, resolved 22.7% wrongly, and left 35.9% needing PET; GRAD resolved 56.2% correctly, 19.5% wrongly, and left 24.3% needing PET (Figure 3B).

The p-Tau181 benchmark was restricted to the 256 A4 gray zone participants with a visit-matched p-Tau181 measurement (55.5% Aβ-positive). In this subset, GRAD Stage 2 achieved an AUC of 0.799 (95% CI, 0.740-0.853), against 0.597 (95% CI, 0.526-0.668) for p-Tau181 (DeLong P < .001) and 0.743 (95% CI, 0.677-0.802) for p-Tau217 alone (P = .06). GRAD’s AUC here (0.799) differs marginally from its AUC across the full external gray zone (0.793, n=630) because this analysis is confined to the p-Tau181 subset; the two are not competing estimates of the same quantity.

Giacomucci et al. report that p-Tau181 correctly reclassified 77.4% (24 of 31) of their indeterminate cases^16^. Applying the same rule - a single dichotomous cutoff derived by Youden’s index, in A4, p-Tau181 classified 59.8% of our gray zone participants correctly and 40.2% incorrectly. GRAD, evaluated on the same 256 participants, classified 58.6% correctly and 21.9% incorrectly while referring the remaining 19.5% for Aβ-PET (Table 2, Figure 3B, C).

**Table 2.**
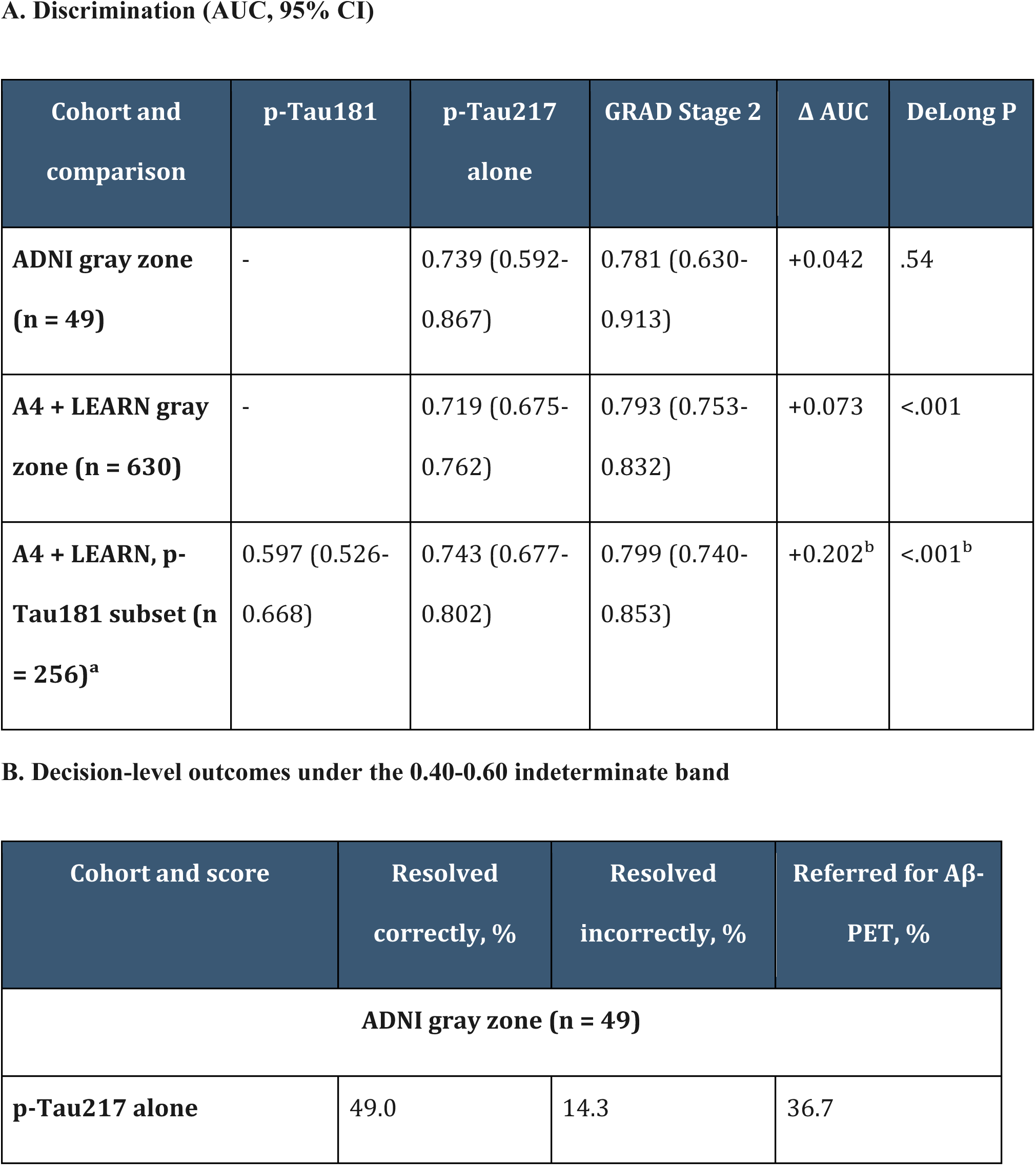
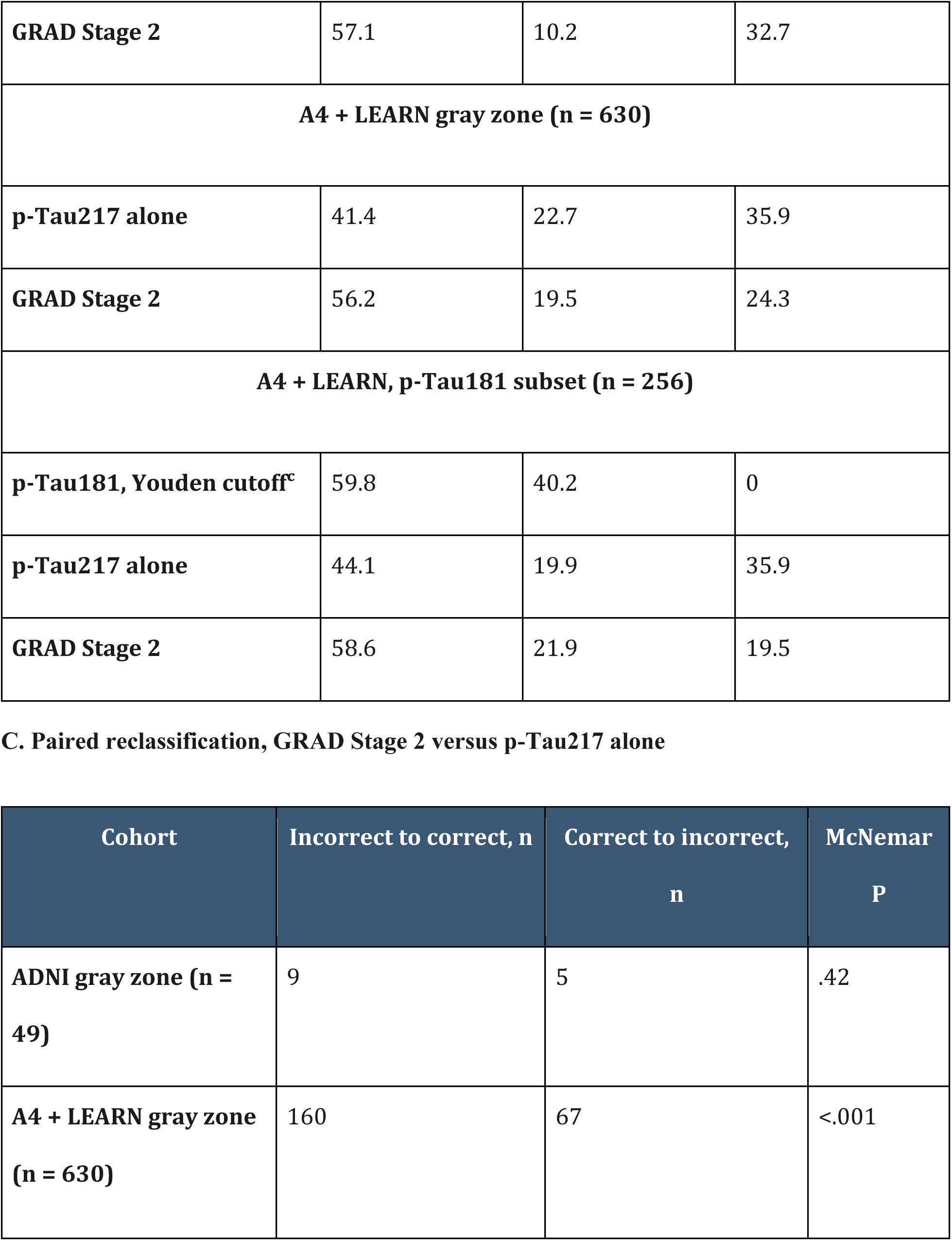
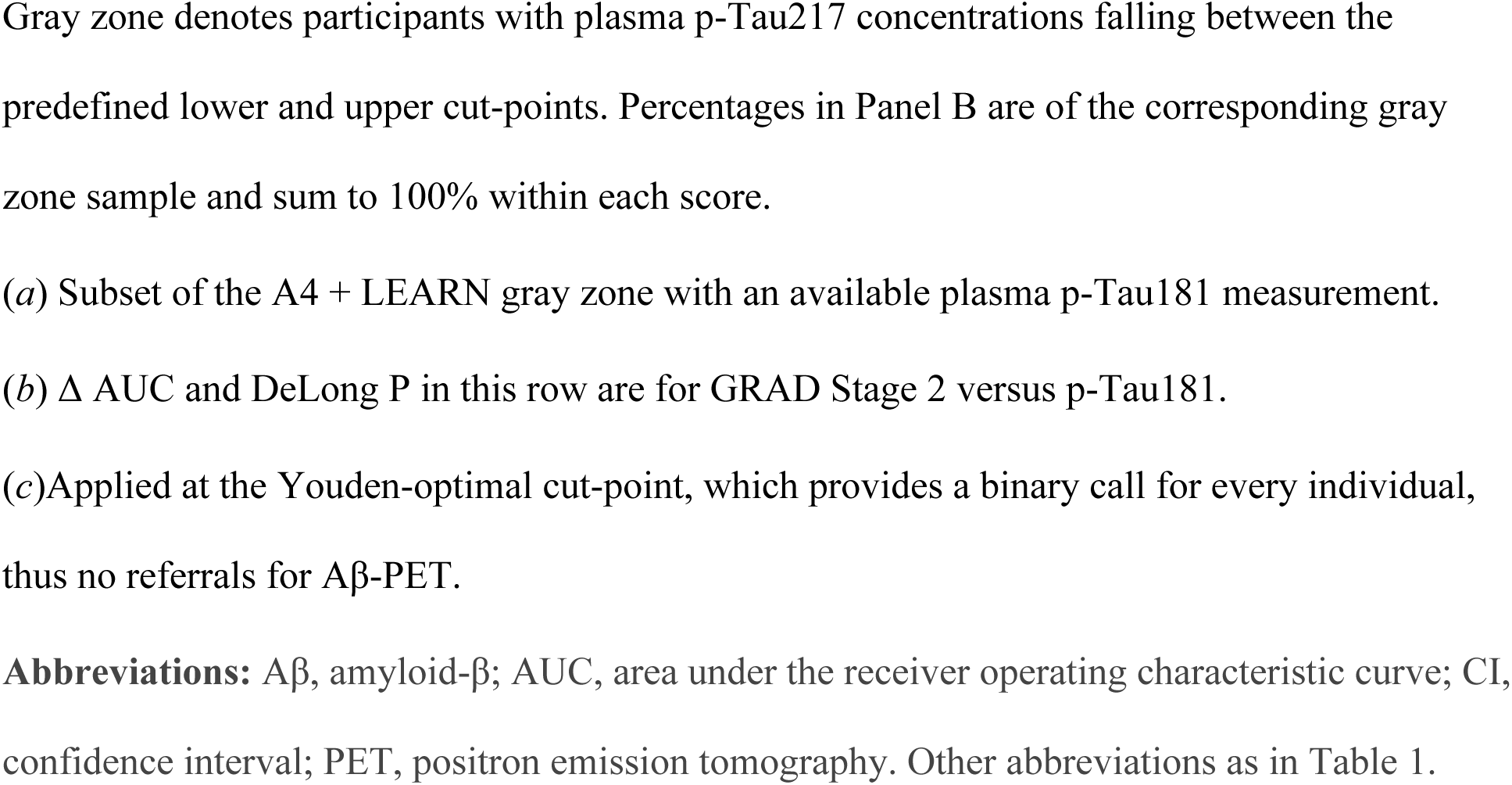
Benchmarking GRAD Stage 2 against p-Tau217 alone and p-Tau181 within the gray zone.

### 3.6 Cost Impact Simulation

Using the external cohort’s routing rates, Universal PET cost $30,000,000 (10,000 scans), p-Tau217 screening with PET for every gray zone case cost $15,000,000 (3,832 scans), a relative 50% saving, and the GRAD staged algorithm cost $7,250,000 (931 scans), a relative 76% saving, or $725 per patient. Using the development cohort’s routing rates the corresponding GRAD figure was $7,655,000 (1,103 scans; 74% saving; $766 per patient). Under development-cohort routing, p-Tau217 screening with PET for every gray zone case cost $13,638,000 (3,379 scans; 55% saving). Measured against this p-Tau217-plus-PET workflow rather than universal imaging, GRAD was 44% less costly under development-cohort routing and 52% less costly under validation-cohort routing. Across both cohorts GRAD reduced PET utilization by 89-91%.

When assessing simulation results as cost per correct diagnosis - counting PET-referred individuals as correctly diagnosed - the external cohort cost was $3,000 for universal PET, $1,639 for p-Tau217 screening with PET for the gray zone, and $863 for GRAD. In the development cohort this was $3,000, $1,423 and $828 respectively (Figure 4).

**Figure 4.**
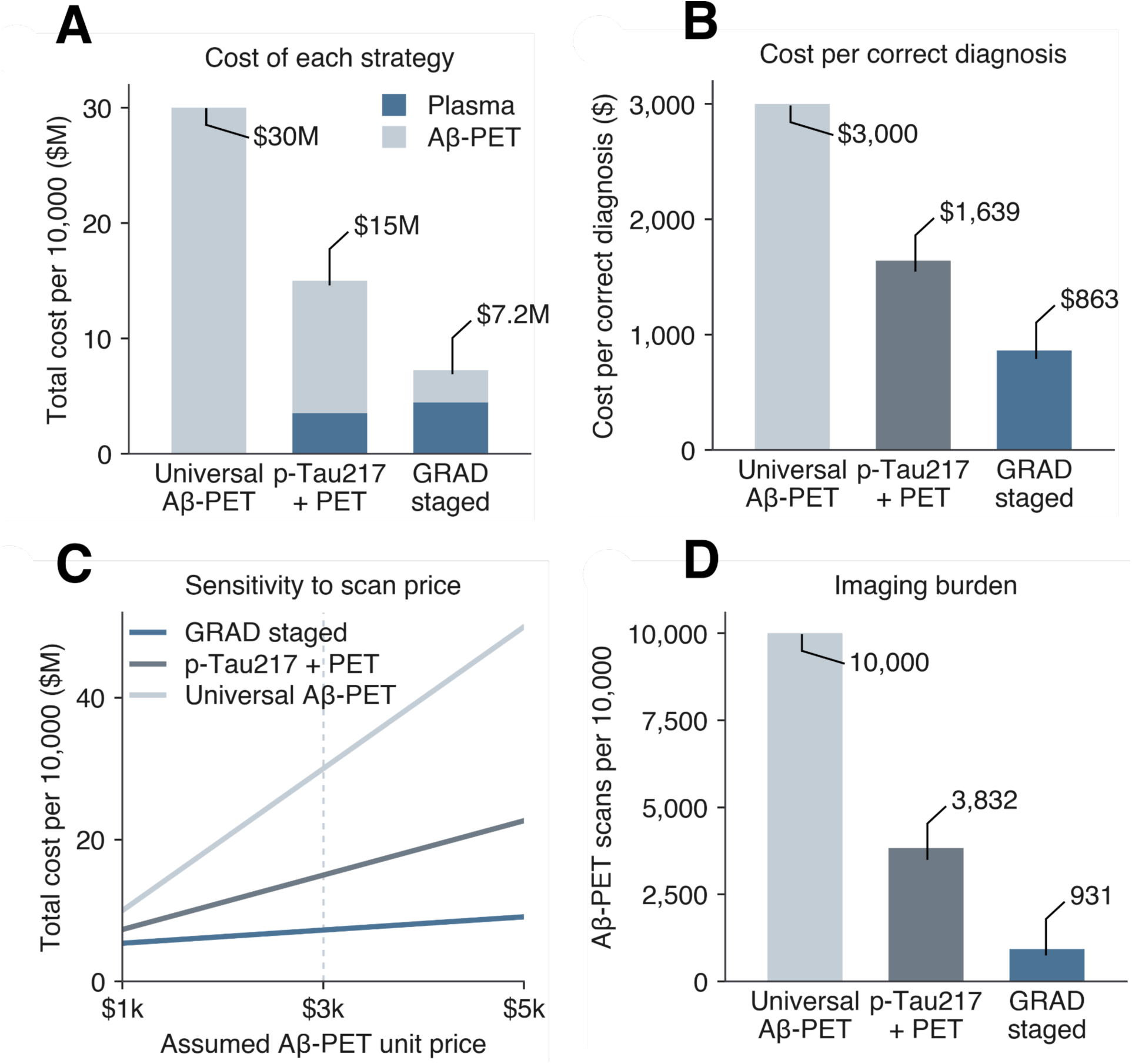
Health Economic Savings Simulation. All panels show a hypothetical cohort of 10,000 patients using external-cohort (A4 + LEARN) routing rates. The unit costs are as follows: $350 per p-Tau217 test, $250 for the Stage 2 multi-analyte panel, and $3,000 per Aβ-PET scan. **(A)** Total cost of each diagnostic strategy, partitioned into plasma assay and the respective Aβ-PET components. **(B)** Cost per correct diagnosis, counting PET-referred patients as correctly diagnosed. **(C)** Sensitivity of total cost to the assumed Aβ-PET unit price across $1,000-$5,000. **(D)** Imaging burden per 10,000 patients, showcasing number of scans per group.

## 4. Discussion

In this multicohort study, we developed and externally validated a two-stage plasma algorithm designed to improve Aβ classification when p-Tau217 is indeterminate. The principal finding was that additional biomarker and genetic information provided meaningful discrimination within this difficult subgroup and transported across two clinically distinct research cohorts. In the A4/LEARN external validation cohort, GRAD Stage 2 improved gray-zone AUC from 0.719 with p-Tau217 alone to 0.793 (P<.001), increased the proportion of participants correctly classified without imaging from 41.4% to 56.2%, and reduced the proportion requiring Aβ-PET from 35.9% to 24.3%. These findings support a staged approach in which p-Tau217 resolves high-confidence cases, additional plasma and genetic information is used selectively for indeterminate results, and confirmatory imaging is retained for residual uncertainty.

Our results also confirm that a substantial p-Tau217 gray zone is present across clinically distinct portions of the AD continuum. A two-cutoff p-Tau217 strategy resolved 96 of 145 cognitively impaired ADNI participants (66.2%) and 1,014 of 1,644 cognitively unimpaired A4/LEARN participants (61.7%), with accuracies of 93.8% and 86.2%, respectively. The remaining gray zone comprised 33.8% of ADNI and 38.3% of A4/LEARN participants, consistent with prior studies reporting that approximately 30-50% of tested individuals fall within an indeterminate range^5, 13–15^. Rather than replacing p-Tau217, GRAD is therefore intended to complement it by concentrating additional testing on the subset for whom the initial biomarker provides insufficient certainty.

Importantly, the principal evidence for GRAD’s incremental value came from external validation. Although Stage 2 improved gray-zone AUC over p-Tau217 alone by 0.042 in ADNI, this difference was not statistically significant in the small development gray zone (n=49; P=.54). In A4/LEARN, however, the improvement was larger (+0.073 AUC; P<.001) despite application to a clinically distinct population, use of a different p-Tau217 assay platform, and scoring without model refitting. GRAD also performed favorably against p-Tau181 integration in the subset with visit-matched measurements, achieving an AUC of 0.799 compared with 0.597 for p-Tau181 (P<.001). Together, these findings suggest that the incremental information provided by the Reflex stage is not confined to the development sample and may offer greater utility for resolving indeterminate p-Tau217 results than reliance on an additional tau marker alone.

Within the Reflex stage, the strongest contributors to gray-zone discrimination were APOE ε4 carrier status, p-Tau217, and the Aβ42/40 ratio. In contrast, GFAP and its interaction with p-Tau217 contributed little additional discrimination, and external ablation analyses similarly showed the largest reductions in AUC when APOE ε4 or Aβ42/40 was removed. This pattern suggests that, once p-Tau217 has identified an indeterminate subgroup, complementary information from amyloid-related measures and genetic risk may be more informative than additional astroglial markers. More broadly, these findings support the rationale for a reflex strategy that combines biologically complementary signals rather than adding biomarkers from the same pathological axis.

The Alzheimer’s Association Clinical Practice Guideline recommends ≥90% sensitivity with ≥75% specificity for a blood-based biomarker used as a triage test, and ≥90% sensitivity and specificity when used in place of Aβ-PET^3^. In the ADNI development cohort, GRAD could be operated at thresholds consistent with the recommended triage benchmark: at the rule-out threshold, sensitivity was 91.8% with 81.7% specificity (NPV 87.5%), while at the rule-in threshold, specificity was 90.0% with 81.2% sensitivity (PPV 92.0%). These operating points were derived in the development cohort, however, and require prospective validation before fixed thresholds can be assumed to meet these benchmarks in other clinical populations. In external validation, the Reflex stage evaluated in isolation achieved 66.7% sensitivity and 80.0% specificity, below the performance recommended for replacement of Aβ-PET. Together, these findings support GRAD as a staged triage and trial-enrichment approach that can reduce reliance on confirmatory imaging while retaining Aβ-PET for residual uncertainty, rather than as a standalone replacement for confirmatory imaging.

The clinical utility of GRAD will depend on the population tested and the respective consequences of false-positive and false-negative classifications. Because predictive values vary with amyloid prevalence, the same operating characteristics yield different projected performance across settings. For example, at an amyloid prevalence of approximately 20%, a negative GRAD result would have a projected NPV of 93.8%, suggesting greatest potential value as a rule-out strategy^7, 46^. At 50% prevalence, projected PPV and NPV are 87.0% and 79.2%, respectively, whereas at 70% prevalence projected PPV increases to 94.0% (Table S9). These estimates illustrate the importance of clinical context but should not be interpreted as prospectively validated performance in primary care, memory clinics, or prevention trials. Thresholds may ultimately require setting-specific calibration depending on amyloid prevalence and the relative consequences of false-positive and false-negative classifications.

A potential implementation strategy would keep p-Tau217 as an initial screen, apply the Reflex multi-analyte panel to individuals with indeterminate results, and then reserve confirmatory Aβ-PET for uncertain cases after Stage 2. In our cohorts, these residual indeterminate cases represented 11.0% of the ADNI development cohort and 9.3% of the A4/LEARN validation cohort. This staged framework could therefore reduce reliance on Aβ-PET while preserving confirmatory imaging for residual uncertainty. Prospective validation in clinically representative populations, including assessment of setting-specific calibration and thresholds, is needed before GRAD is used to guide individual clinical or trial-enrollment decisions.

Utilizing GRAD’s diagnostic strategy carries potential economic and access implications. In our simulation, GRAD reduced projected testing costs by 74-76% relative to universal PET and by 44-52% relative to p-Tau217 screening with PET confirmation of the gray zone, with estimated costs of $725-766 per patient. GRAD also had the lowest modeled cost per correct diagnosis ($828-863), although these estimates do not incorporate the downstream consequences or costs of misclassification. Blood-based testing generally requires less specialized infrastructure than Aβ-PET, although the multi-analyte panel required for GRAD’s Reflex stage is not yet routinely available in all settings. By reducing projected PET requirements by 89-91%, this staged approach could be particularly useful in settings where access to specialized neuroimaging is limited, including rural and underserved communities where dementia diagnoses are often less timely and complete^47, 48^.

### 4.1 Limitations

Our study also has several limitations. First, both cohorts are predominantly white (89% of ADNI and 94% of A4) and highly educated, so GRAD’s performance in racially and ethnically diverse populations is unknown and may differ^30, 49^. Second, our study also excludes renal function, a known confounder of plasma biomarker concentrations^55–57^, along with any comorbid proteinopathies^4, 19^ and mixed pathologies^38^. Third, our development cohort is small, and its gray zone contains only 49 participants. The improvement in ADNI Stage 2 did not reach significance on its own (P = .54) - we therefore rest GRAD’s benefit on the well-powered external cohort; the internal estimate is reported as supportive. Fourth, our Stage 2 required a multi-analyte panel that is not yet routine in all settings. Also, analyte availability was uneven: Aβ42/40 was measured in 919 of 1,644 A4 participants, and missing values were imputed from training-set medians - an imperfect measure. The analysis benchmarking GRAD against p-Tau181 also carries limitations. It was possible only in A4, because ADNI’s plasma p-Tau181 was assayed a median of four years before the p-Tau217 draws used here and only 7 of 49 ADNI gray zone participants have a visit-matched value. We suggest the comparison should be read as a single-cohort comparison between populations that differ (impaired versus unimpaired). Our cross-sectional design limits any assessment of performance over time or during treatment monitoring. Fifth, although discrimination transferred across cohorts, calibration did not fully transport, indicating that predicted probabilities and fixed probability thresholds may require recalibration before application in populations with different amyloid prevalence or assay characteristics. Finally, our cost simulation projects savings hypothetically rather than with real-world data. It ignores geographic variation, and does not price the cost of misclassification, including ARIA. Real cohort financial data would be required to demonstrate the economic case directly.

### 4.2 Conclusion

As p-Tau217 is increasingly incorporated into early AD screening, GRAD offers a staged approach for addressing the substantial subgroup with indeterminate results. In external validation, the Reflex stage improved discrimination over p-Tau217 alone within the gray zone while retaining Aβ-PET for residual uncertainty. Across the full pipeline, 89.0% of ADNI and 90.7% of A4/LEARN participants received an Aβ classification without imaging, suggesting that selective reflex testing could substantially reduce reliance on confirmatory PET. Modeled testing costs were also 44-52% lower than strategies relying on PET for all indeterminate p-Tau217 results, although these projections require confirmation with real-world economic data. At present, GRAD should be viewed as a potential triage and trial-enrichment strategy rather than a replacement for confirmatory imaging. Future work should prospectively evaluate the algorithm in clinically heterogeneous and racially and ethnically diverse populations, assess setting-specific calibration and thresholds, and determine whether the projected reductions in PET use and testing costs are realized in clinical practice.

## Author Contributions

H.P.: Conceptualization, Methodology, Software, Formal Analysis, Investigation, Data Curation,

Writing - Original Draft, Project Administration.

C.V.: Validation, Writing-Review & Editing.

C.B.: Methodology, Supervision, Review & Editing

E.K.: Formal Analysis, Investigation, Data Curation

C.U.: Visualization, Writing-Original Draft.

A.B.: Supervision, Resources, Writing-Review & Editing.

B.F.: Conceptualization, Supervision, Project Administration, Writing-Review & Editing.

## Acknowledgments

We thank all individuals participating in the ADNI, A4 and LEARN Trial Cohort(s). The A4 Study was a secondary prevention trial in preclinical Alzheimer’s disease, aiming to slow cognitive decline associated with brain amyloid accumulation in clinically normal older individuals. The A4 Study was funded by a public-private-philanthropic partnership, including funding from the National Institutes of Health-National Institute on Aging, Eli Lilly and Company, Alzheimer’s Association, Accelerating Medicines Partnership, GHR Foundation, an anonymous foundation, and additional private donors, with in-kind support from Avid Radiopharmaceuticals, Cogstate, Albert Einstein College of Medicine and the Foundation for Neurologic Diseases.The companion observational Longitudinal Evaluation of Amyloid Risk and Neurodegeneration (LEARN) Study was funded by the Alzheimer’s Association and GHR Foundation. The A4 and LEARN Studies were led by Dr. Reisa Sperling at Brigham and Women’s Hospital, Harvard Medical School, and Dr. Paul Aisen at the Alzheimer’s Therapeutic Research Institute (ATRI) at the University of Southern California. The A4 and LEARN Studies were coordinated by ATRI at the University of Southern California, and the data are made available by the Alzheimer’s Clinical Trial Consortium through the Global Research & Imaging Platform (GRIP). The complete A4 Study Team list is available on: https://www.actcinfo.org/a4-study-team-lists/. We would like to acknowledge the dedication of the study participants and their study partners who made the A4 and LEARN Studies possible.

## Ethics Statement

ADNI was approved by the institutional review boards of all participating institutions, and the A4 and LEARN Studies were approved by the institutional review board at each participating site. All participants provided written informed consent prior to enrollment. Our study was not prospectively registered.

## Data Availability Statement

All code for our analysis is available at: https://github.com/hsparankusham/GRAD-grayzone-classifier.

The ADNI datasets analyzed during this study are available from the ADNI repository at https://adni.loni.usc.edu, and the A4/LEARN datasets are available through the Alzheimer’s Clinical Trial Consortium’s Global Research & Imaging Platform at https://www.actcinfo.org, both subject to registered data use agreements.

