## Supplements: Tables S1-S9, Figure S1 for "A Two-Stage Plasma Biomarker Algorithm Reduces Amyloid PET Use and the p-Tau217 Gray Zone"

**The PDF file includes:**

Tables S1 to S9

Figure S1

**Supplementary Table S1.** Analyte availability by cohort and assay platform.

| **Cohort and assay platform** | **n** | **p-Tau217** | **GFAP** | **Aβ42/40** | **APOE genotype** |
|---|---|---|---|---|---|
| ADNI - Janssen immunoassay | 64 | 64 | 0 | 0 | 64 |
| ADNI - Lumipulse G (Fujirebio, UPENN) | 81 | 81 | 81 | 80 | 68 |
| ADNI - all | 145 | 145 | 81 | 80 | 132 |
| A4 + LEARN - Lilly MSD / Roche Elecsys | 1,644 | 1,644 | 931 | 919 | 1,642 |

Counts are participants with a non-missing value for each analyte. ADNI p-Tau217 was measured on the Lumipulse G assay (Fujirebio, University of Pennsylvania) and on the Janssen assay; the Janssen subset contributed p-Tau217 only. A4 + LEARN p-Tau217 was measured on the Lilly Research Laboratories MSD immunoassay, with GFAP and Aβ42/40 on the Roche Elecsys platform in a subset of the cohort. ADNI counts refer to the 145 cognitively impaired participants who formed the development cohort. Because the two cohorts used different p-Tau217 platforms, all plasma biomarkers were reference-anchored within the cohort and platform before analysis (Section 2.3).

**Supplementary Table S2.** Complete-panel sensitivity analysis in A4 + LEARN.

| A4 + LEARN subgroup | n | Aβ+, % | Full pipeline AUC (95% CI) | Gray zone AUC | Accuracy, % |
|---|---|---|---|---|---|
| All participants | 1,644 | 69.6 | 0.879 (0.861-0.895) | 0.793 | 80.3 |
| Complete panel | 915 | 55.7 | 0.854 (0.828-0.878) | 0.754 | 79.3 |
| Imputed ≥1 analyte | 729 | 87.1 | 0.888 (0.852-0.918) | 0.765 | 81.5 |

Participants are those in the external validation cohort with a non-missing value for all seven Stage 2 features. The ADNI-fitted model is unchanged; only the participants scored differ. Panel availability was informative: Aβ prevalence was 55.7% among complete-panel participants against 87.1% among those requiring imputation, so the two subgroups are not interchangeable and the difference in AUC between them (-0.034; 95% CI, -0.075 to +0.008) reflects case mix as well as any effect of imputation. The interval does not exclude a difference of clinical relevance and should not be read as demonstrating equivalence.

**Supplementary Table S3.** Sensitivity of external validation to the reference-anchoring definition.

| A4 + LEARN reference definition | Uses Aβ status | Stage 1 AUC | Gray zone, % | Gray zone AUC | Full pipeline AUC | Accuracy, % |
|---|---|---|---|---|---|---|
| Aβ-negative participants (primary) | Yes | 0.867 | 38.3 | 0.793 | 0.879 | 80.3 |
| Lowest p-Tau217 tercile | No | 0.867 | 21.2 | 0.708 | 0.874 | 82.4 |
| Whole cohort, median and IQR | No | 0.867 | 42.1 | 0.787 | 0.871 | 66.3 |
| Whole cohort, mean and SD | No | 0.867 | 34.4 | 0.748 | 0.867 | 58.2 |
| Random half of Aβ-negative participants | Yes | 0.867 | 37.2 (31.6-42.5) | 0.790 (0.777-0.808) | 0.880 (0.878-0.881) | 80.4 (78.8-82.2) |

Because our reference population was defined by Aβ-status, the transformation we applied to the validation cohort was estimated using that cohort's outcome labels. We thus repeated the external validation with the A4 reference redefined, without reference to Aβ-status, as the lowest tertile of the p-Tau217 distribution (n=548, 33.9% of whom were actually Aβ-positive) (Table S3).

The ADNI-fitted model is held completely fixed; only the mu and sigma used to reference-anchor the validation cohort change. Stage 1 AUC is computed over the whole cohort and is identical to four decimal places under every definition, because the Stage 1 probability is a monotone function of raw p-Tau217 whatever reference is chosen. The final row draws the reference from a random half of the Aβ-negative participants, repeated 200 times, and reports the median with the 2.5th to 97.5th percentile range. The two whole-cohort definitions deliberately mis-anchor the scale, since 69.6% of the cohort is Aβ-positive, and are included to bound the worst case: discrimination is preserved, while the 0.5 probability threshold is displaced and accuracy falls accordingly. AUC, area under the receiver operating characteristic curve.

**Supplementary Table S4.** Cost model unit costs and assumptions.

| Cost component | Unit cost | Applied to |
| --- | --- | --- |
| Plasma p-Tau217, single analyte | $350 | Every patient, except under universal Aβ-PET |
| Multi-analyte reflex add-on | $250 | Only patients routed to Stage 2 |
| Aβ-PET scan | $3,000 | Universal strategy: every patient. Staged strategies: indeterminate cases only |
| Simulated cohort size | 10,000 | All strategies |

Unit costs were drawn from 2024 U.S. Medicare clinical laboratory and physician fee schedules (references 43-45). Where a range was plausible we adopted the higher plasma figure and a conservative Aβ-PET price so that savings are not overstated. Costs are undiscounted, expressed in 2024 U.S. dollars, and exclude clinician time, downstream treatment, and the consequences of misclassification.

**Supplementary Table S5.** Diagnostic performance of GRAD by stage and cohort.

| Cohort and stage | n | AUC (95% CI) | Sensitivity, % | Specificity, % | PPV, % | NPV, % | LR+ | LR- | Brier |
|---|---|---|---|---|---|---|---|---|---|
| ADNI, Stage 1 | 96 | 0.939 (0.866-0.995) | 94.9 | 91.9 | 94.9 | 91.9 | 11.71 | 0.06 | - |
| ADNI, Stage 2 | 49 | 0.781 (0.630-0.913) | 76.9 | 73.9 | 76.9 | 73.9 | 2.95 | 0.31 | 0.191 |
| ADNI, full pipeline | 145 | 0.915 (0.859-0.961) | 89.4 | 85.0 | 89.4 | 85.0 | 5.96 | 0.12 | 0.108 |
| A4 + LEARN, Stage 1 | 1,014 | 0.904 (0.884-0.923) | 82.8 | 94.1 | 97.0 | 70.1 | 13.99 | 0.18 | - |
| A4 + LEARN, Stage 2 | 630 | 0.793 (0.753-0.832) | 66.7 | 80.0 | 88.1 | 51.8 | 3.33 | 0.42 | 0.201 |
| A4 + LEARN, full pipeline | 1,644 | 0.879 (0.861-0.895) | 76.7 | 88.6 | 93.9 | 62.3 | 6.71 | 0.26 | 0.145 |

Stage 1 rows cover only the participants that stage resolves; its sensitivity and specificity use the 0.25 and 0.75 routing thresholds as the decision rule. Stage 2 rows cover the gray zone. Stage 2 and full-pipeline values use a 0.5 probability threshold. Confidence intervals are percentile bootstrap over participants (2,000 resamples). ADNI values are leave-one-out cross-validated; A4 + LEARN values are from the ADNI-fitted model applied without refitting. AUC, area under the receiver operating characteristic curve; LR, likelihood ratio; NPV, negative predictive value; PPV, positive predictive value.

**Supplementary Table S6.** Operating-point sweep in the ADNI development cohort.

| Operating point | Probability threshold | Sensitivity, % | Specificity, % | PPV, % | NPV, % | LR+ | LR- | Accuracy, % |
|---|---|---|---|---|---|---|---|---|
| Rule-out (90% sensitivity target) | 0.451 | 91.8 | 81.7 | 87.6 | 87.5 | 5.01 | 0.10 | 87.6 |
| Rule-in (90% specificity target) | 0.667 | 81.2 | 90.0 | 92.0 | 77.1 | 8.12 | 0.21 | 84.8 |

Both operating points are derived in the ADNI development cohort (n = 145) from leave-one-out cross-validated predictions of the full pipeline, and are reported to show the range over which the model can be tuned. They have not been validated prospectively and should not be adopted as fixed thresholds in other populations without recalibration. LR, likelihood ratio; NPV, negative predictive value; PPV, positive predictive value.

**Supplementary Table S7.** Stage 2 associations and model coefficients.

| Feature | Univariable OR per SD (95% CI) | P | Multivariable OR per SD (95% CI) | Penalised coefficient |
|---|---|---|---|---|
| APOE ε4 carrier | 3.14 (1.53-6.44) | .002 | 2.62 (0.92-7.51) | +0.442 |
| p-Tau217 | 2.34 (1.20-4.56) | .013 | 8.16 (1.10-60.54) | +0.344 |
| Aβ42/40 | 0.32 (0.13-0.77) | .011 | 0.16 (0.02-1.06) | -0.329 |
| tau-Aβ42/40 ratio | 1.80 (0.94-3.47) | .078 | 0.12 (0.01-1.16) | +0.072 |
| Age | 0.58 (0.31-1.10) | .097 | 0.40 (0.12-1.35) | -0.205 |
| GFAP | 0.80 (0.43-1.48) | .482 | 0.41 (0.05-3.07) | -0.104 |
| GFAP × p-Tau217 | 0.94 (0.53-1.67) | .829 | 2.50 (0.30-20.59) | -0.042 |

All estimates are from the ADNI gray zone (n = 49). Univariable odds ratios fit each feature alone and are the values quoted in Section 3.4. Multivariable odds ratios fit all seven features together without penalisation and are reported so that the two can be compared directly; their intervals are wide because the features are correlated and the sample is small. The final column gives the L2-penalised coefficients of the deployed Stage 2 model on standardised features, whose magnitudes are plotted in Figure 2E. The negative coefficient for Aβ42/40 reflects that a lower ratio indicates greater amyloid burden. The univariable estimates and the penalised coefficients agree in ranking APOE ε4, p-Tau217 and Aβ42/40 highest. The unpenalised multivariable estimates are unstable, with p-Tau217 and the tau-Aβ42/40 ratio taking large coefficients of opposing sign because the two are collinear by construction; this instability is why the deployed model is L2-penalised. OR, odds ratio.

**Supplementary Table S8.** Feature ablation in the A4 + LEARN gray zone.

| Model | Gray zone AUC | Δ AUC vs full model |
|---|---|---|
| Full seven-feature model | 0.793 | - |
| Without APOE ε4 carrier | 0.758 | -0.034 |
| Without Aβ42/40 | 0.763 | -0.030 |
| Without p-Tau217 | 0.773 | -0.020 |
| Without tau-Aβ42/40 ratio | 0.792 | -0.001 |
| Without GFAP × p-Tau217 | 0.793 | +0.000 |
| Without Age | 0.797 | +0.004 |
| Without GFAP | 0.800 | +0.008 |
| Plus NfL | 0.769 | -0.023 |

Each row refits Stage 2 on the ADNI gray zone with the stated feature set and scores the A4 + LEARN gray zone (n = 630) without refitting. A negative Δ AUC indicates that removing the feature degraded external discrimination. NfL was evaluated as an eighth feature and excluded on this basis.

**Supplementary Table S9.** Predictive values across assumed Aβ prevalence.

| Assumed Aβ prevalence, % | PPV, % | NPV, % |
|---|---|---|
| 20 | 62.7 | 93.8 |
| 30 | 74.2 | 89.9 |
| 50 | 87.0 | 79.2 |
| 70 | 94.0 | 61.9 |

Positive and negative predictive value depend on how common Aβ-positivity is in the population being tested, whereas sensitivity and specificity do not. Values are derived from the full-pipeline operating point in A4 + LEARN (76.7% sensitivity, 88.6% specificity) applied at each assumed prevalence by Bayes theorem. The 30% and 50% rows approximate a primary care memory complaint population and a specialist memory clinic respectively; the observed prevalence was 69.6% in A4 + LEARN and 58.6% in ADNI. NPV, negative predictive value; PPV, positive predictive value.

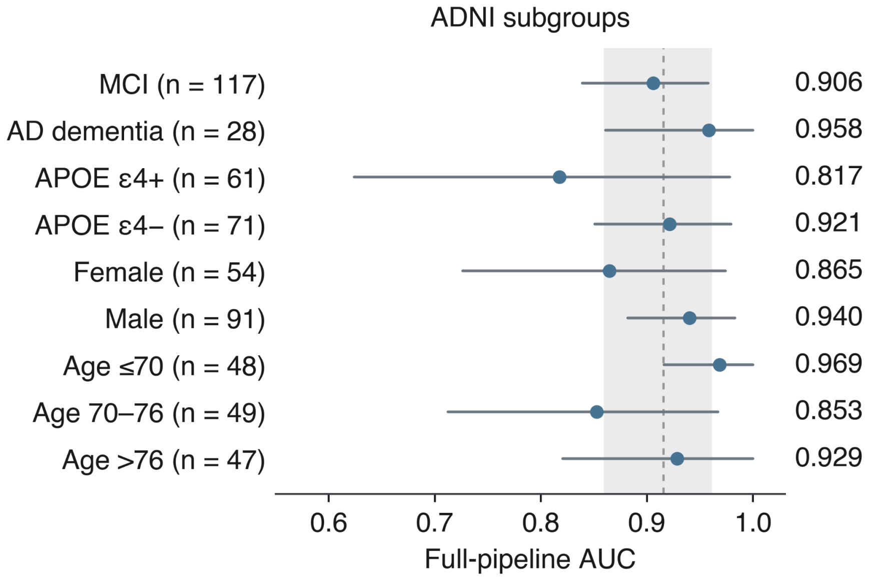


**Supplementary Figure S1.** Full-pipeline discrimination by subgroup, ADNI development cohort. Area under the receiver operating characteristic curve for the complete GRAD pipeline within clinical subgroups of the ADNI development cohort (n = 145), with 95% percentile bootstrap confidence intervals. The dashed line and shaded band show the whole-cohort estimate and its interval. Intervals are wide for the smaller strata, notably AD dementia (n = 28) and APOE ε4 carriers (n = 61), and should be read as imprecision rather than instability. The corresponding analysis in the A4 + LEARN validation cohort is Figure 3A; cognitive status cannot be stratified there because all participants are cognitively unimpaired.
